# Neonatal resuscitation: an observational study assessing the readiness of service providers in Nepal

**DOI:** 10.1101/2022.10.20.22281310

**Authors:** Robert B Clark, Mala Chalise, Ranjan P Dhungana

## Abstract

A significant proportion of neonatal mortality, a major public health challenge in low- and middle-income countries, can be attributed to intrapartum-related hypoxic events. This study seeks to assess the determinants of health care providers’ competence in providing newborn resuscitation. A cross-sectional survey of 154 health facilities in Nepal was done. This nested study evaluated the newborn resuscitation knowledge and skills of 462 health care providers by individually assessing a delivery using a 22-item clinical practice observation tool and administering the standard 18-item Helping Babies Breathe Knowledge Check

Significant predictors of provider knowledge included: province (0.085 points higher in Bagmati province, p=0.018); mean availability of essential utilities and resuscitation aids (0.173 points, p<0.001 and 0.187, p= 0.02 respectively); participation in – Latter-day Saint Charities – Safa Sunaulo Nepal (LDSC/SSN) newborn resuscitation training, scale-up and skill retention program (0.676 units higher, p<0.001); and qualifications (0.136 points higher among health providers with Bachelor of Nursing degree, p<0.001, 0.072 points higher among providers with Masters in Nursing degree, p= 0.010 and 0.110 units higher among providers with Senior Auxiliary Nursing Midwife degree,, p=0.001).

Significant factors associated with resuscitation skill included province (0.056 units higher in province 1, p= 0.015 and 0.037 units higher in Sudurpaschim province, p=0.034); delivery caseload (0.066 units higher mean skill score in health facilities with average monthly delivery of more than 120, p= 0.011); availability of newborn resuscitation practice aids (0.093 units higher score in health facilities with resuscitation practice aids, p= 0.008); and participation in LDSC/SSN newborn resuscitation training, scale-up and skill retention program (0.968 units, p< 0.001).

Participation in the LDSC/SSN’ skill retention program was the best predictor of newborn resuscitation knowledge and skills. The LDSC/SSN model of newborn resuscitation training, scale up and skill retention could be one potential cost-effective model to address gaps in resuscitation knowledge and skills among service providers.

## Introduction

Neonatal mortality, newborn death between birth and 28 days, is a significant public health challenge in low-and middle-income countries(1,2). Of the 2.4 million neonatal deaths that occurred globally in 2020, nearly 43% occurred in Sub-Saharan Africa and 36% in Central and South Asia. Globally, nearly half of the under-five mortality occurs during the first 28 days of life; mainly due to preterm birth, intrapartum-related complications such as birth asphyxia, and infections(3). Intrapartum-related Hypoxic Events (IHE), also referred to as birth asphyxia, accounts for nearly 30 to 35 percent of neonatal deaths worldwide(4).

The world has witnessed phenomenal progress in reductions of the under-five mortality rate in the past few decades. However, the reductions in neonatal mortality have lagged, resulting to an increased proportion of overall child deaths attributed to the neonatal period(3). Newborn deaths are projected to account for nearly 52% of child deaths by 2030(5). The first 24 hours of birth holds the greatest risk for neonatal mortality and represents the period of greatest need for high-quality newborn care(6).

A similar trend in the proportion of neonatal deaths attributable to child deaths has been observed in Nepal, where the reduction in neonatal mortality has fallen far behind the under-five mortality rate(7). IHE, resulting in the failure to initiate and sustain a normal process of breathing at birth, continues to be one of the leading causes of neonatal mortality in low-income countries, including Nepal(8,9).

Evidence suggests that many of these deaths can be prevented by ensuring quality antenatal care, improved intrapartum care, skilled birth attendance at delivery, and emergency obstetric care(10). Neonatal resuscitation, a globally accepted toolset for managing perinatal asphyxia and/or mitigating its long-term negative consequences, includes interventions at the time of birth to support establishment of breathing and circulation(11). It is estimated that approximately 5-10% of newborns require simple stimulation and/or suctioning within the first minute (the “Golden Minute”) after birth, 3-6% require assisted ventilation, and less than 1% require advanced resuscitation to help them breathe(12). Facility-based neonatal resuscitation, including timely and appropriate assisted ventilation, can prevent intrapartum-related neonatal deaths by 30%(13).

In response to the lack of resuscitation knowledge and skills among birth attendants in resource-limited settings, the American Academy of Pediatrics developed Helping Babies Breathe (HBB) in 2010(14). This training package was introduced in Nepal in 2012. Currently, three Ministry of Health approved in-service curricula in Nepal, the Community-Based Integrated Management of Neonatal and Childhood Illness (CB-IMNCI), the SBA training package, and the HBB/HMS Training Course utilize HBB materials. HBB has been supported by multiple child health partners in Nepal, including UNICEF. The National Health Training Center (NHTC), a nodal agency of the Nepal Ministry of Health and Population (MOHP) for disseminating approved in-service provider education throughout the country, has orchestrated HBB training-of-trainer (TOT) courses in Nepal since 2014 in collaboration with Latter-day Saint Charities (LDSC) and Safa Sunaulo Nepal (SSN).

However, training in resuscitation knowledge and skills alone is not sufficient for the accelerated decline in neonatal mortality required to achieve Sustainable Development Goals targets. Efforts to improve perinatal outcomes must also focus on improving the Quality of Care (QoC) for newborns(15). WHO’s Quality of Care Framework places enormous emphasis on safety, effectiveness, timeliness, efficiency, and an equitable and people-centered care approach to maternal and newborn care services(16). According to WHO, QoC during childbirth in health facilities depends on availability of physical infrastructure, supplies, and competent human resources with the knowledge, skills, and capacity to provide pregnancy and childbirth services(17).

It is estimated that high quality maternal health services could lead to the prevention of 1 million newborn deaths(18). The readiness of health facilities to provide newborn care through necessary infrastructure, equipment, drugs and skilled human resources is a necessary first step in improving QoC (19,20). Although improving the quality of care in health facilities is considered increasingly indispensable to end preventable mortality and morbidity among newborns(17), there is a critical gap in defining the determinants of quality newborn care, including skills assessment of service providers.

This study seeks to assess the determinants of health care providers’ competence in providing newborn resuscitation. The results of this study can assist in identification of health provider QoC in the treatment and management of asphyxiated babies, care gaps, and strengthening provider competence. This study may also be helpful in offering recommendations for future efforts in reducing intrapartum-related neonatal mortality in Nepal.

## Methods

### Ethical Approval

Ethical approval from the study was approved from the Nepal Health Research Council with registration number 236/2022 P.

### Study Design

We conducted a cross-sectional survey of health facilities in private and various levels of public hospitals from all seven provinces of Nepal to capture current readiness status for immediate newborn care. Readiness was measured in terms of the availability of essential utilities, equipment, drugs, medical supply, resuscitation aids, and staff competency to provide immediate newborn care. This nested study reports on the status and determinants of provider competency in newborn resuscitation.

### Sample Size

The sample size of health facilities to be included in the study was calculated with the following formula(21):

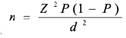

where, n is the sample size, Z is the statistic corresponding to 95% level of confidence, P is the expected prevalence and d is precision. This study used the prevalence of newborn resuscitation to calculate the desired sample size. Considering that 10% of the newborns require resuscitation within the first minute after birth(12), 5% precision(22) and non-response rate of 13%, a sample size was estimated at 156 for the study. Health facilities with average annual delivery caseload of 400, were purposely selected to be included in the survey. Of the sample, 154 facilities responded and were therefore included in the study.

To assess providers’ resuscitation skills, three deliveries per facility were observed for a total of 462 deliveries. The 462 health care providers who assisted the deliveries completed the knowledge assessment.

### Data Collection

Data were collected by two experienced HBB-trained health personnel. To investigate providers’ competency to offer newborn resuscitation, the data collector directly observed and assessed three deliveries per facility, using a 22-item clinical practice observation tool based on the HBB algorithm(23). A summary of the key skills assessed is presented in table 1. Each correctly performed skill was rated as 1 while partially correct or incorrect steps were rated as 0.

**Table 1.** Summary of key skills observed.

| **S.No.** | **Items Observed** |
| --- | --- |
| 1. | **Prepares for the birth** |
|  | Identifies helper and review emergency plan |
|  | Prepares the area for delivery room |
|  | Performs handwash |
|  | Tests the function of bag and mask and suction device |
| 2. | **Performs routine care** |
|  | Dries baby thoroughly |
|  | Recognizes crying |
|  | Keeps baby warm |
|  | Checks breathing |
|  | Clamps or ties and cuts the umbilical cord |
|  | Positions on mother's chest to encourage breastfeeding |
| 3. | **Clear Airways and stimulation** |
|  | Positions airway in neutral position |
|  | If meconium and secretions present, lightly suctions the mouth and nose |
|  | Stimulates the baby by rubbing the back if the baby is not breathing |
|  | Assesses breathing (looks, listens and feels) |
| 4. | **Bag and Mask Ventilation** |
|  | Correctly identifies need for assisted ventilation (if needed) |
|  | Correctly positions baby head in extension |
|  | Evaluates proper use of bag and mask, including use of appropriate bag and mask size, seal and chest movement |
|  | Initiates ventilation within Golden Minute |
|  | Provides assisted ventilation correctly (30-50 bpm) |
|  | Evaluates for breathing or chest movement |
|  | Continues ventilation |
|  | Plans for advanced care, if required |

Next, a knowledge assessment was conducted with each of the three service providers per facility using the standard HBB Knowledge Check. This 18-item multiple choice questionnaire assesses various domains of newborn resuscitation(24), with each correct answer rated 1 while incorrect answers were rated as 0. Pretesting of the knowledge and skill assessment tools were done in Sindhupalchok and Banke districts.

This study uses data on facility readiness to provide immediate newborn care measured in five domains: availability of essential utilities, equipment, drugs, medical supply and resuscitation aids. Details on data collection method, tools, performance standards, and measurement methods for each domain are reported elsewhere(25).

Permission from hospital administrations and the Nepal Health Research Council was obtained before data collection. Verbal consent was also obtained from the service providers prior to observation of deliveries and HBB knowledge check. The completeness of collected data were regularly checked by the data collectors before leaving each health facility, throughout the entire data collection period.

### Measurements

The newborn resuscitation knowledge and skill scores are the outcome variables in this study. Skill scores and knowledge scores were created as a composite score by adding the presence of each item in a 22-item clinical practice observation tool and 18-item scale knowledge check tool, respectively. Equal weightage was given to each item-scale in both the tools. The score of both skill observations and knowledge test were converted to 100%. Health care providers whose knowledge score was 75% or higher were considered having adequate knowledge to provide newborn resuscitation. Health care providers whose skill score was 100% were considered skillful in providing newborn resuscitation.

The outcome variables were examined against the set of factors considered to be contributing to provider competence (Fig 1). These are the potential explanatory variables that may influence a provider’s resuscitation knowledge and skill.

**Figure 1.**
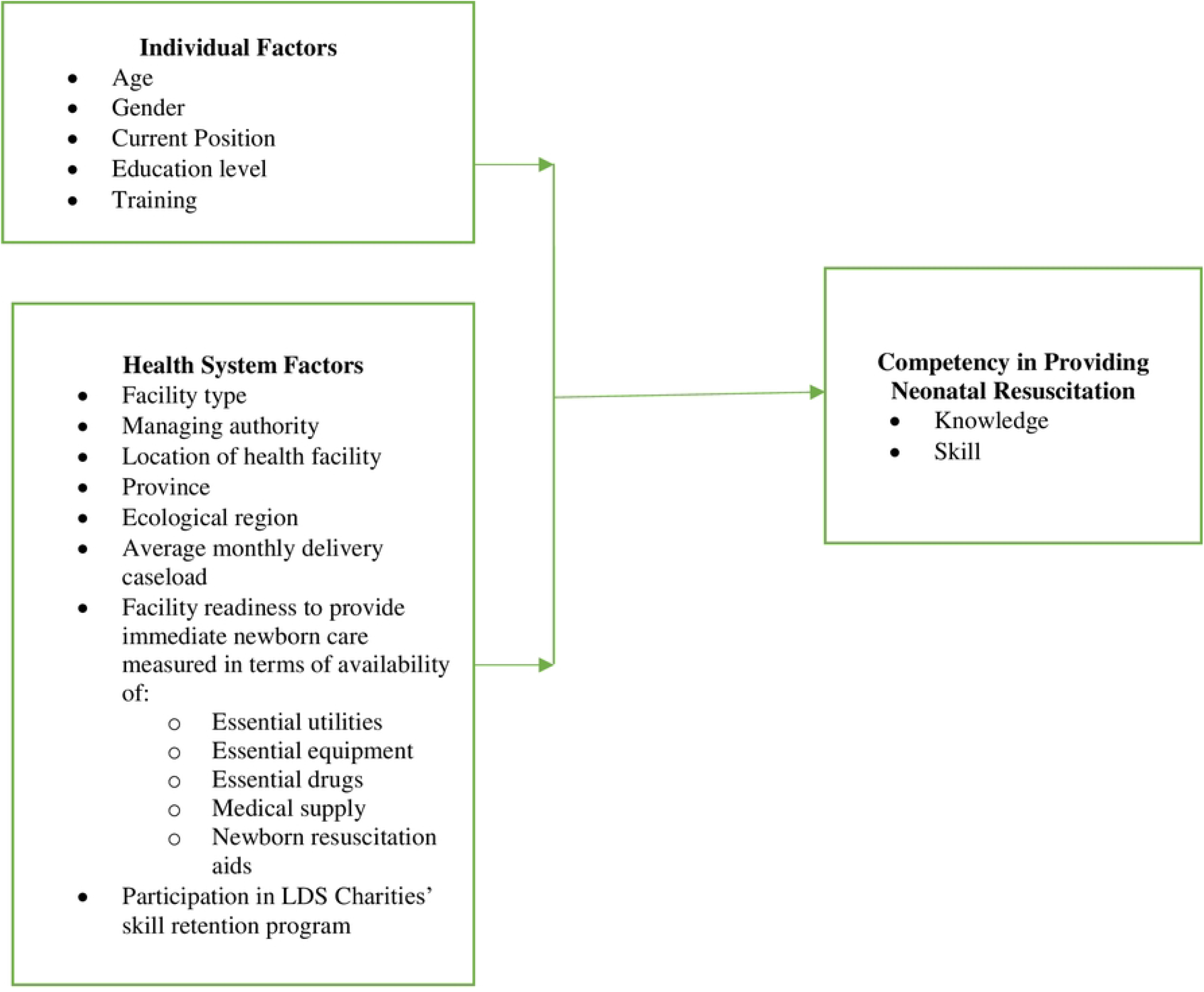
The conceptual framework.

### Data Management and Analysis

The survey data collected on paper were exported to Excel and was cleaned for accuracy, completeness and consistency. The Excel data was imported to SPSS version 26, where further statistical analysis was performed.

Descriptive analyses were used to summarize health facility and provider characteristics. A composite skill score and knowledge score were created as previously described. Linear regression analysis was performed to identify associations between different predictive variables and outcome variables, i.e., the neonatal resuscitation knowledge and skills score. Pearson’s correlation coefficient of more than 0.7 was used to check for multicollinearity among the independent variables. Multicollinearity was observed between the independent variables ‘training’ and ‘facility support of LDSC/SSN’. Similarly, multicollinearity was observed between the independent variables ‘facility readiness index to provide immediate newborn care’, and ‘essential utilities score’. Therefore, variables ‘training’ and ‘facility readiness index to provide immediate newborn care’ were removed from the final models. A p value of less than 0.05 was considered statistically significant association.

## Results

### Characteristics of health facilities

Table 2 presents a summary of the general characteristics of the facilities included in the study. Out of 154 facilities involved in the study, more than half were in urban areas (55.2%) and the majority were Local level facilities (51.3%). Most of the facilities were managed by the government (82.5%) and most were in the Terai region (66.9%). The average number of deliveries per facility per month was 124, with nearly half (44.2%) delivering sixty or less babies per month. Half of the health facilities under the study had LDSC/SSN intensive support for HBB training, scale up, and skill maintenance.

**Table 2.**
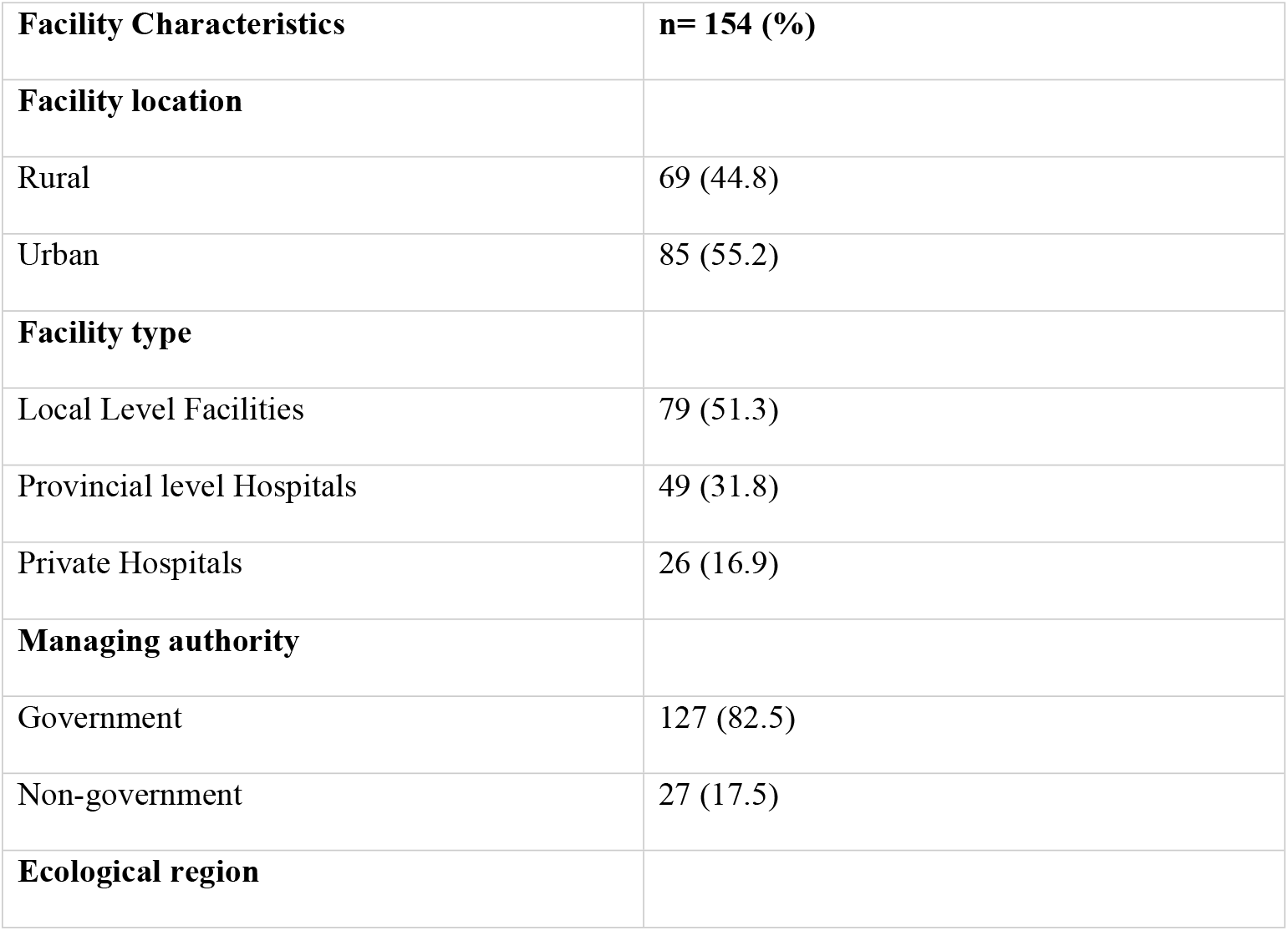
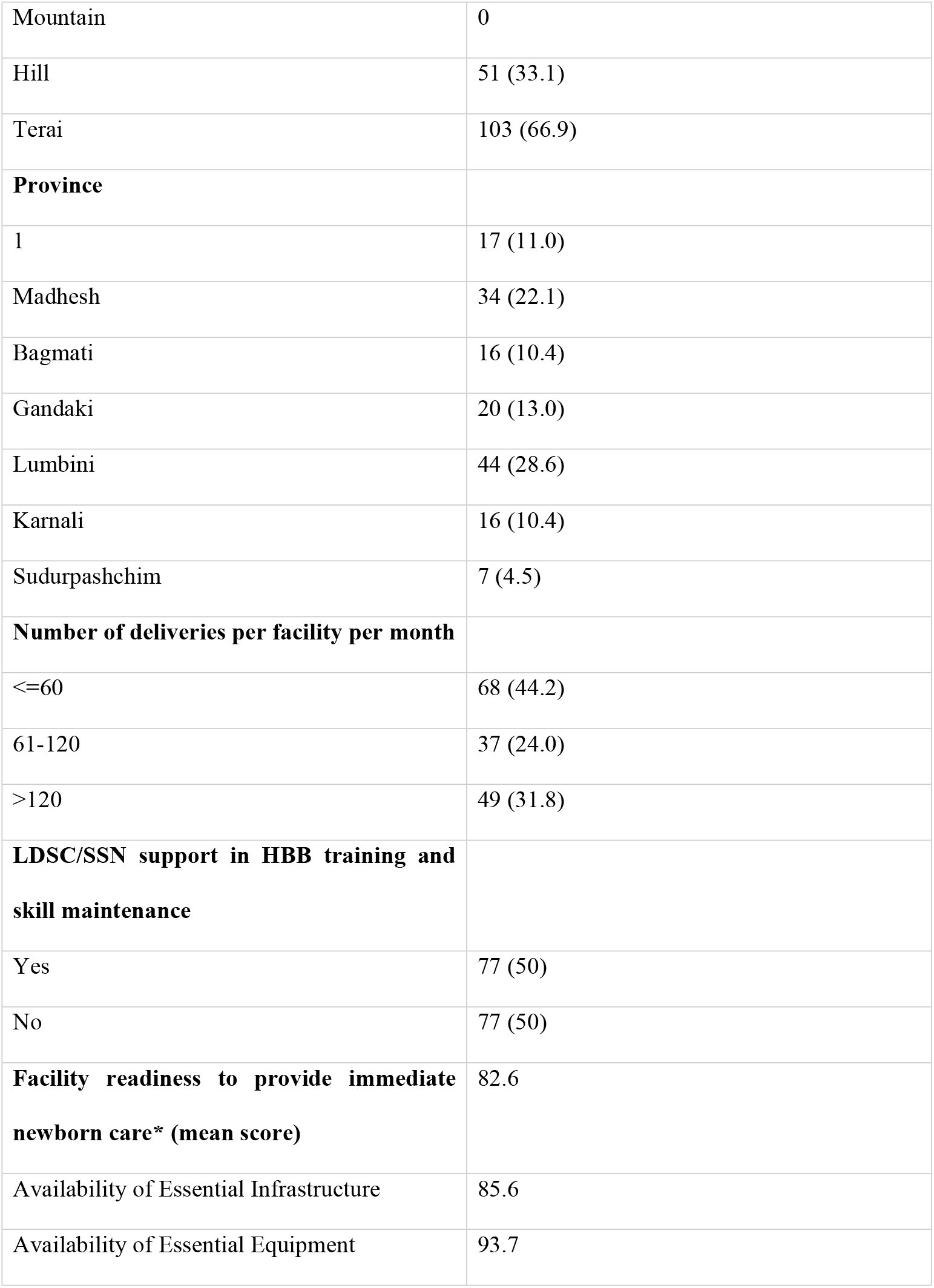
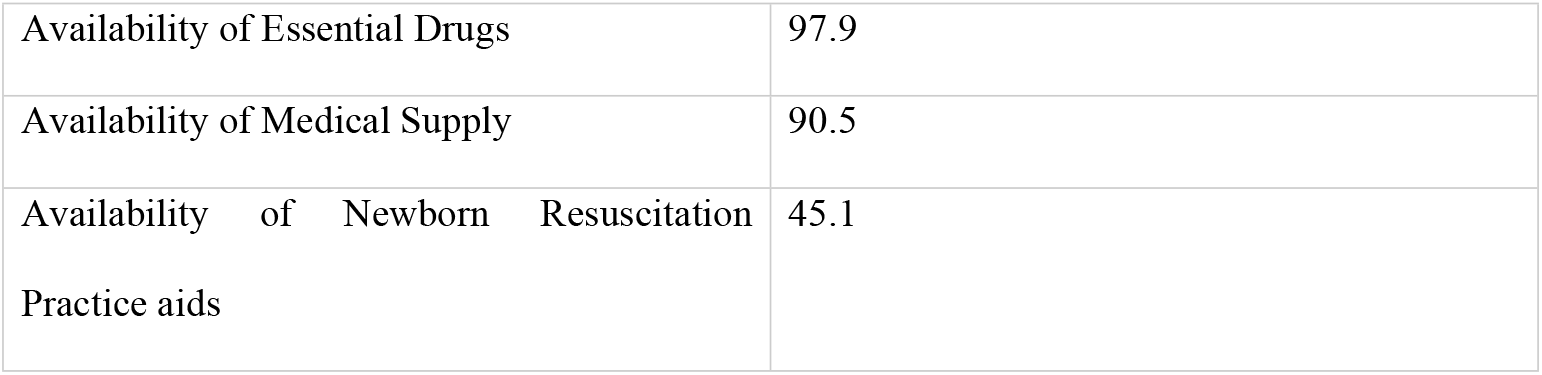
Characteristics of health facilities.

| **Facility Characteristics** | **n= 154 (%)** |
| --- | --- |
| **Facility location** |  |
| Rural | 69 (44.8) |
| Urban | 85 (55.2) |
| **Facility type** |  |
| Local Level Facilities | 79 (51.3) |
| Provincial level Hospitals | 49 (31.8) |
| Private Hospitals | 26 (16.9) |
| **Managing authority** |  |
| Government | 127 (82.5) |
| Non-government | 27 (17.5) |
| **Ecological region** |  |
| Mountain | 0 |
| Hill | 51 (33.1) |
| Terai | 103 (66.9) |
| **Province** |  |
| 1 | 17 (11.0) |
| Madhesh | 34 (22.1) |
| Bagmati | 16 (10.4) |
| Gandaki | 20 (13.0) |
| Lumbini | 44 (28.6) |
| Karnali | 16 (10.4) |
| Sudurpashchim | 7 (4.5) |
| **Number of deliveries per facility per month** |  |
| <=60 | 68 (44.2) |
| 61-120 | 37 (24.0) |
| >120 | 49 (31.8) |
| **LDSC/SSN support in HBB training and skill maintenance** |  |
| Yes | 77 (50) |
| No | 77 (50) |
| **Facility readiness to provide immediate newborn care\* (mean score)** | 82.6 |
| Availability of Essential Infrastructure | 85.6 |
| Availability of Essential Equipment | 93.7 |
| Availability of Essential Drugs | 97.9 |
| Availability of Medical Supply | 90.5 |
| Availability of Newborn Resuscitation Practice aids | 45.1 |

Each facility received a score for readiness to perform immediate newborn care, derived from the five domains listed in Table 2. The overall facility readiness index to provide immediate newborn care was around 83%.

The domains of essential utilities, essential equipment, essential drugs, and medical supplies had an average readiness score of more than 85%, while the availability of resuscitation aids had the average readiness score of about 45%. Fig 2 provides distribution of health facilities by immediate newborn care readiness score and LDS Charities support. Details of facility readiness to provide immediate newborn care including lists of tracer items and performance standards for the five domains of facility readiness are reported elsewhere(25).

**Figure 2.**
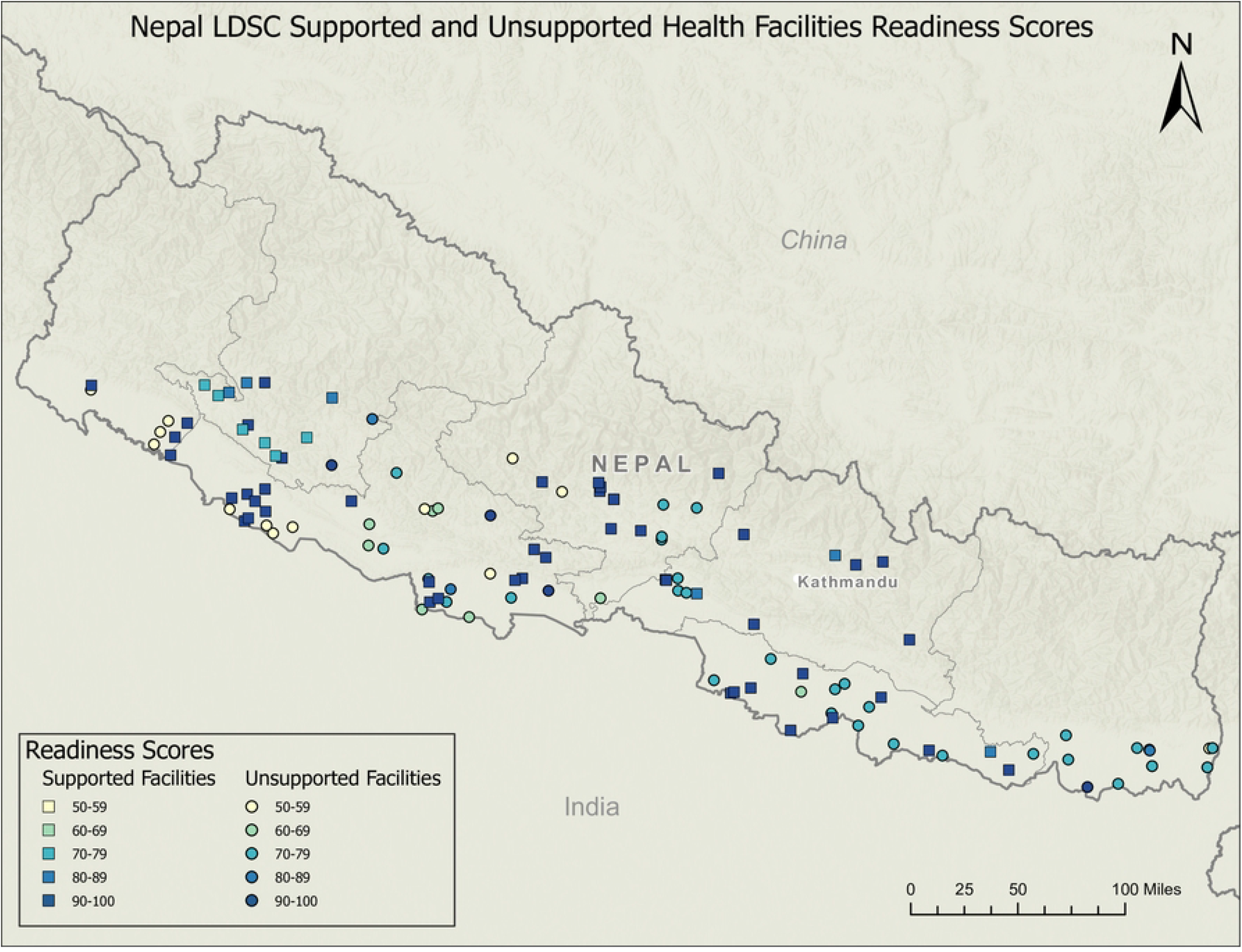
Distribution of health facilities by readiness score and LDS Charities support.

### Characteristics of health care providers

Table 3 presents the general characteristics of the 462 health care providers included in the study. The majority of providers were in the age group 20-29 years. All of the providers were female and majority of them were Staff Nurse (SN) (51.7%). The majority had previously attended either Skilled Birth Attendant (SBA) training (50%) or both HBB and SBA training (49.4%).

**Table 3.** Characteristics of health care providers.

| **Individual Characteristics** | **n (%)** |
| --- | --- |
| **Age** | Mean age= 32.3 (SD= 7.74) |
| 20-29 | 228 (49.4) |
| 30-39 | 143 (31) |
| 40-49 | 64 (13.9) |
| 50-59 | 27 (5.8) |
| **Gender** |  |
| Female | 462 (100) |
| Male | 0 (0) |
| Years of experience in current position |  |
| **Education Level** |  |
| Auxiliary Nurse Midwife (ANM) | 69 (14.9) |
| Bachelor of Nursing (BN) | 69 (14.9) |
| BSc Nursing | 1 (0.2) |
| Masters of Nursing (MN) | 3 (0.6) |
| Senior ANM (SANM) | 81 (17.5) |
| Staff Nurse (SN) | 239 (51.7) |
| **Training** |  |
| Helping Babies Breathe (HBB) | 3 (0.6) |
| Skilled Birth Attendant (SBA) | 231 (50) |
| Both HBB and SBA | 228 (49.4) |

### Providers’ knowledge and skill on newborn resuscitation

Table 4 depicts shows the mean resuscitation knowledge score of providers was slightly higher (66.7) than the mean skill score (60.0). Mean knowledge and skill scores of providers were higher in health facilities from urban areas (84.0and 72.4 respectively). Likewise, health care providers from Provincial level hospitals had higher mean knowledge (86.0) and skill (85.6) scores as compared to those from Local level and private hospitals. Providers from government managed health facilities had higher mean knowledge (82.0) and skill (62.4) scores. Service providers from hill region had higher knowledge and skills scores (84.2 and 78, respectively). Knowledge score was higher among health care providers from Bagmati province (85.2) while skill score was higher among service providers from Gandaki province (82.5).

**Table 4.** Knowledge and skill score by background characteristics.

| Facility Characteristics | Number of observation (n) | Knowledge | Skill |
| --- | --- | --- | --- |
|  |  | Mean (SD) | Mean (SD) |
| **Overall** | 462 | 66.7 (7.88) | 60.0 (43.02) |
| **Facility location** |  |  |  |
| Rural | 207 | 79.2 (5.89) | 44.4 (42.939) |
| Urban | 255 | 84.0 (8.61) | 72.4 (38.758) |
| **Facility type** |  |  |  |
| Local Level Facilities | 237 | 79.6 (6.75) | 48.5 (42.861) |
| Provincial level Hospitals | 147 | 86.0 (7.99) | 85.6 (28.533) |
| Private Hospitals | 78 | 80.8 (7.84) | 46.8 (45.470) |
| **Managing authority** |  |  |  |
| Government | 381 | 82.0 (7.89) | 62.4 (42.101) |
| Non-government | 81 | 81.7 (7.84) | 48.8 (45.741) |
| **Ecological region** |  |  |  |
| Mountain | - | - | - |
| Hill | 153 | 84.2 (7.21) | 78.0 (35.953) |
| Terai | 309 | 80.7 (7.95) | 51.1 (43.510) |
| **Province** |  |  |  |
| 1 | 51 | 74.8 (4.49) | 33.5 (29.930) |
| Madhesh | 102 | 80.8 (7.84) | 44.5 (48.556) |
| Bagmati | 48 | 85.2 (8.60) | 69.2 (46.196) |
| Gandaki | 60 | 84.1 (7.89) | 82.5 (28.855) |
| Lumbini | 132 | 82.2 (7.81) | 58.5 (41.231) |
| Karnali | 48 | 83.6 (5.95) | 79.3 (37.228) |
| Sudurpashchim | 21 | 83.9 (6.78) | 79.8 (24.519) |
| **Number of deliveries per facility per month** |  |  |  |
| <=60 | 204 | 80.3 (6.48) | 52.9 (43.604) |
| 61-120 | 111 | 82.0 (7.81) | 59.0 (44.318) |
| >120 | 147 | 83.9 (9.17) | 70.7 (39.176) |
| **LDSC/SSN support in HBB training and skill maintenance** |  |  |  |
| Yes | 231 | 87.9 (5.70) | 99.1 (4.581) |
| No | 231 | 75.7 (4.11) | 20.9 (24.853) |
| **Education Level** |  |  |  |
| Auxiliary Nurse Midwife (ANM) | 69 | 78.5 (6.73) | 43.0 (40.388) |
| Bachelor of Nursing (BN) | 69 | 88.9 (7.76) | 89.2 (28.179) |
| BSc Nursing | 1 |  |  |
| Masters of Nursing (MN) | 3 | 90.7 (8.49) | 66.7 (57.735) |
| Senior ANM (SANM) | 81 | 81.1 (4.56) | 46.1 (43.881) |
| Staff Nurse (SN) | 239 | 80.9 (7.91) | 61.1 (42.558) |
| **Training** |  |  |  |
| Helping Babies Breathe (HBB) | 3 | 85.2 (3.21) | 100 (0.00) |
| Skilled Birth Attendant (SBA) | 231 | 75.7 (4.11) | 20.9 (24.853) |
| Both HBB and SBA | 228 | 88.0 (5.72) | 99.1 (4.610) |
| **Age of the service providers** |  |  |  |
| 20-29 | 228 | 81.3 (7.79) | 58.9 (42.333) |
| 30-39 | 143 | 81.4 (7.70) | 55.7 (43.702) |
| 40-49 | 64 | 84.6 (8.27) | 70.7 (43.416) |
| 50-59 | 27 | 82.5 (5.91) | 67.6 (41.495) |

Service providers from health facilities with average monthly deliveries of more than 120 had higher knowledge (83.9) and skill (70.7) scores. Similarly, service providers from facilities that had participated in LDSC/SSN program of newborn resuscitation training, skill maintenance and scale-up had higher knowledge and skill scores (87.9 and 99.1, respectively). While service providers with Masters of Nursing (MN) degree had higher knowledge score (90.7), the skill score was higher among service providers with Bachelor of Nursing (BN) degree (89.2). Newborn resuscitation knowledge score was higher among providers with both HBB and SBA training (88.0), while skill score was highest among service providers with HBB training (100). Providers from age group 40-49 years had higher mean knowledge (84.6) and skill (70.7) scores as compared to other age groups, while providers from age group 20-29 had lowest knowledge score (81.2) and providers from 30-39 age group had lowest skill score (55.7).

Overall, 78.6% (n= 362) of health care providers had a knowledge score of 75% or more and were therefore considered having adequate knowledge to provide newborn resuscitation. In contrast, only 48.3% (n= 223) of providers had a skill score of 100% and were thus considered competent to provide newborn resuscitation. Detailed findings on each item in a 22-item clinical practice observation, and each item in an 18-item scale knowledge check tool is provided in supplementary figure and table (S1 and S2, respectively).

### Factors associated with newborn resuscitation knowledge and skill score

Table 5 shows the results of multiple linear regression analysis for newborn resuscitation knowledge and skill score. The findings depict that 66.7% (Adjusted R Square= 0.667, *P*<0.001) of the variance in newborn resuscitation knowledge score and 88.7% of the variance in skill score (Adjusted R Square= 0.887, *P*<0.001) can be accounted for by the predictors collectively, in both models respectively.

**Table 5:** Multiple linear regression analysis of newborn resuscitation knowledge score and skill score.

| Facility Characteristics | Knowledge |  | Skill |  |
| --- | --- | --- | --- | --- |
|  | Standardized Coefficient | p-value | Standardized Coefficient | p-value |
| **Facility location** |  |  |  |  |
| Rural | Ref |  | Ref |  |
| Urban | 0.066 | 0.175 | 0.014 | 0.611 |
| **Facility type** |  |  |  |  |
| Local Level Facilities | Ref |  | Ref |  |
| Provincial level Hospitals | 0.049 | 0.262 | 0.006 | 0.800 |
| Private Hospitals | -0.049 | 0.749 | 0.43 | 0.630 |
| **Managing authority** |  |  |  |  |
| Government | Ref | - | Ref | - |
| Non-government | 0.087 | 0.561 | -0.099 | 0.288 |
| **Ecological region** |  |  |  |  |
| Terai | Ref |  | Ref |  |
| Hill | -0.041 | 0.394 | 0.003 | 0.917 |
| **Province** |  |  |  |  |
| 1 | -0.067 | 0.117 | 0.056 | **0.015** |
| Madhesh | Ref | - | -0.088 | **<0.001** |
| Bagmati | 0.085 | **0.018** | -0.022 | 0.264 |
| Gandaki | 0.029 | 0.548 | 0.040 | 0.104 |
| Lumbini | 0.072 | 0.089 | Ref | - |
| Karnali | 0.006 | 0.899 | 0.021 | 0.426 |
| Sudurpaschim | 0.025 | 0.433 | 0.037 | **0.034** |
| **Number of deliveries per facility per month** |  |  |  |  |
| <=60 | Ref |  | Ref |  |
| 61-120 | 0.017 | 0.648 | -0.012 | 0.592 |
| >120 | 0.037 | 0.393 | 0.066 | **0.011** |
| **Facility readiness to provide immediate newborn care** |  |  |  |  |
| Availability of Essential Utilities | 0.173 | **<0.001** | -0.056 | 0.052 |
| Availability of Essential Equipment | -0.056 | 0.230 | -0.009 | 0.737 |
| Availability of Essential Drugs | 0.053 | 0.118 | 0.095 | 0.000 |
| Availability of Medical Supply | -0.035 | 0.498 | -0.214 | 0.000 |
| Availability of Newborn Resuscitation Practice aids | 0.187 | **0.002** | 0.093 | **0.008** |
| **LDSC/SSN support in HBB training and skill maintenance** |  |  |  |  |
| Yes | 0.676 | <**0.001** | 0.968 | <**0.001** |
| No | Ref | - | Ref | - |
| **Education Level** |  |  |  |  |
| Auxiliary Nurse Midwife (ANM) | 0.020 | 0.518 | -0.005 | 0.800 |
| Bachelor of Nursing (BN) | 0.136 | <**0.001** | -0.018 | 0.340 |
| BSc Nursing | -0.023 | 0.410 | -0.012 | 0.468 |
| Masters of Nursing (MN) | 0.072 | **0.010** | -0.016 | 0.318 |
| Senior ANM | 0.110 | **0.001** | -0.037 | 0.054 |
| Staff Nurse (SN) | Ref | - | Ref | - |
| **Age of the service providers** |  |  |  |  |
| 20-29 | Ref | - | Ref | - |
| 30-39 | -0.024 | 0.426 | 0.003 | 0.862 |
| 40-49 | 0.002 | 0.959 | -0.008 | 0.634 |
| 50-59 | -0.043 | 0.152 | 0.010 | 0.570 |
| **Knowledge score** | - | - | -0.013 | 0.646 |

Results reveal that health care providers from Bagmati province had a 0.085 units higher mean knowledge score (p=0.018) as compared to health care providers from Madhesh province. Similarly, service providers from health facilities that have higher mean availability of essential utilities and resuscitation aids have a higher knowledge score (0.173 points, p<0.001 and 0.187, p= 0.02 respectively). Results also depict that health care providers from health facilities with LDSC/SSN support in newborn resuscitation training, skill maintenance and scale-up have a 0.676 units higher mean knowledge score (p<0.001) than of those health care providers from health facilities with no LDSC/SSN support in newborn resuscitation. Similarly, the mean knowledge score was higher for health care providers with qualifications such as BN (0.136points, p<0.001), MN (0.072 points, p= 0.010) and Senior Auxiliary Nurse Midwife (SANM)(0.110 units, p=0.001) than those providers whose qualification was SN.

Findings from multiple linear regression of newborn resuscitation skill score shows that health care providers from province 1 and Sudurpaschim Province have 0.056 units (p= 0.015) and 0.037 units (p=0.034) higher mean skill score as compared to providers from Lumbini province.

In contrast, providers from Madhesh province have 0.088 units (p=0.000) lower newborn resuscitation skill score as compared to health care providers from Lumbini province. Health care providers from those health facilities with average monthly delivery of more than 120 also have 0.066 units higher mean skill score (p= 0.011) than those providers from health facilities with average delivery per month of less than 60. Like the knowledge score, the skill score was also higher among health care providers from health facilities with higher mean availability of resuscitation aids (0.093 units, p= 0.008) and from health facilities with LDSC/SSN support (0.968 units, p< 0.001).

## Discussion

This study assessed correlates of provider newborn resuscitation competency, measured in terms of knowledge and skill. This is one of the most recent studies to assess provider knowledge and skill in newborn resuscitation in Nepal. Direct observation of actual resuscitations in the clinical setting to assess provider skill and use of a standardized tool to measure resuscitation knowledge are the notable strengths of this study. Inclusion of a variety of health facilities, from basic Local level to advanced Provincial level facilities, representing all seven provinces, is also a strength of this study. This study may therefore offer a generalizable and comprehensive analysis of correlates of resuscitation knowledge and skill at different levels of clinical settings in Nepal.

Our study observed a slight knowledge-skill gap (66.67% vs 60.3%) in newborn resuscitation. A low concordance between knowledge and skill found in this study is consistent with a study in India, though the difference was much higher compared to our study (76% vs. 24%)(26). The observed discrepancy between knowledge and skill may be in part due to a lack of sufficient practice-either through attendance of deliveries or manikin practice - to maintain clinical competency in resuscitation. A Skill Birth Attendant (SBA) training (a 3-months in-service training program targeted to nurses, doctors and midwives to enhance their competency for safe birth, including management of normal deliveries and complications) follow up study in Nepal found a huge knowledge-skill gap owing to sub-optimal utilization of skills learned through attendance of deliveries below the WHO recommended threshold required to maintain clinical competency(27).

The SBA training utilizes an abbreviated form of the HBB curriculum for management of asphyxiated babies (27), resulting into poor exposure to newborn resuscitation and skill acquisition (Written communication, Dr. Robert B Clark, September 2, 2022.) This may have led to low skill score despite majority of service providers receiving prior SBA training.

In 2018, LDSC and SSN jointly designed and administered a newborn resuscitation training, scale-up and skill retention program, funded by LDSC. It centered on developing and maintaining the capacity of facility-based trainers to sustain the skills required to manage newborn emergencies. The program utilized evidence-based strategies to implement a cost-effective program for HBB scale up, skill retention, and support. The program prepared facility-based HBB trainers and supplied them with training materials, skill practice manikins (NeoNatalies), and basic delivery room clinical equipment. Facility-based trainers took the lead in scaling up and sustaining the skills through on-the-job cascade trainings to physicians, nurses/midwives in their respective facilities, encouraging regular practice sessions, and refresher trainings. An External Mentor (one per portfolio of hospitals) used evidence-based strategies for supporting these trainers. The strategies included regular monitoring visits to review practice logs; conducting practice drills and Bag and Mask Skill Checks; weekly telephone meetings with facility-based trainers; and monthly in-person meetings with hospital clinical leadership to facilitate training and utilization of skills. The finding of higher skill score compared to knowledge score among the facilities that participated in LDSC/SSN program indicates the role of ongoing practice in sustaining skill. Health facilities that participated in LDSC/SSN program of newborn resuscitation training, scale-up and skill retention had greater practice opportunities through low-dose high-frequency (LDHF) practice of bag and mask ventilation the NeoNatalie simulation device combined with frequent refresher training and mentorship. Participation in the LDSC/SSN program the strongest predictor of both knowledge (beta Coeff=0.676, p<0.001) and skill (beta Coeff=0.968, p<0.001).

The LDSC/SSN’ model of newborn resuscitation training, scale-up and skill retention has been implemented in selected health facilities from six provinces of Nepal (except province 1). With facility-based trainers training the service providers in their facilities, the program has led to scale-up of newborn resuscitation skills by cascading the HBB training to 231 medical personnel in Madhesh Province, 711 medical personnel in Gandaki Province, 1,785 providers in Lumbini Province. The program has also been linked to reduction in key perinatal outcomes. Studies have suggested the supportive role of the program in reduction in neonatal deaths under 24 hours of life (p<0.001) and intrapartum stillbirths (p<0.001)(28)(29). The positive association between LDSC/SSN support to health facilities and knowledge and skill scores observed in this study speaks to the importance of emphasizing on various QI measures, besides initial training, in acquiring, sustaining and scaling-up of newborn resuscitation competency. Interestingly, the time at which retention activities with an external LDSC/SSN mentor were transitioned to internal quality retention efforts varied widely. The LDSC/SSN scale up and skill retention efforts utilizing an external mentor included four portfolios of facilities, concluding in 2019 (two portfolios), 2020, and 2021.

Our study also adds to the evidence that utilization of newborn resuscitation practice aids such as NeoNatalie and newborn resuscitation guidelines/ algorithms have a positive influence on both knowledge and skill of the service providers. In a setting with low delivery caseload, and therefore limited opportunities for resuscitation, frequent refresher training, mentorships and practicing with anatomical models are effective strategies to sustain skill over time.(30) Nonetheless, since the majority of the health facilities under study were deficient in the practice aids, this should be urgently addressed to provide a conducive environment for skill maintenance.

Surprisingly, our analysis revealed that availability of essential utilities such as availability of a functional newborn care corner, 24 hours availability of delivery staff, and so on was one of the predictors of knowledge score (p<0.001) among health workers but had no effect on providers’ skill. Our findings also point out that other domains of facility readiness such as availability of essential medical equipment, drugs and supplies are not as critical for improved knowledge and skill of service providers in newborn resuscitation.

Unlike another study(31), this study found no correlation between newborn resuscitation knowledge and skill scores, thus indicating the relative inability of health care providers to translate their knowledge into skill. The clinician’s ability to translate newborn resuscitation knowledge into skill requires rapid assessment and judgement, prompt decision making and immediate action(32) all of which comes from ongoing practice and utilization of learned skills.

Our findings indicate that the monthly delivery caseload is positively associated with providers’ skill (p= 0.011) but not with providers’ knowledge of resuscitation. Health facilities with higher deliveries are more likely to manage non-breathing infants than those with fewer deliveries, thus making health workers more accustomed in performing resuscitation. This further establishes the importance of practice in acquiring and retaining clinical competency.

Our study found that providers with higher qualifications such as BN, MN and SANM were positively associated with knowledge score only, implying that providers’ education level has bearings on their knowledge but not their resuscitation skill. This might be due to the fact that health workers with high degrees such as SANM, BN and MN could have better exposure to evidence-based newborn resuscitation information than other providers. Our finding of association between provider’s educational level and knowledge score is consistent with other studies from low-income countries where nurses and midwives with Bachelor and higher-level nursing degree had greater knowledge on newborn resuscitation(33).

This study revealed higher knowledge and skill scores among providers working at Provincial level hospitals as compared to Local level facilities and private hospitals. In contrast, a cross-sectional study that assessed newborn resuscitation knowledge and skills of health workers at different levels of health facilities in Afghanistan noted a difference in knowledge score but not skill scores between health care providers working at various levels of health facilities(34). Overall, our findings suggest that providers in Provincial hospitals had better competency to provide resuscitation than providers from the other health facilities. This might be due to both the high volume of delivery in Provincial level hospitals and referral of complicated cases, thus requiring the health workers to resuscitate newborns more frequently than in other types of health facilities.

Both knowledge and skill scores were higher among health workers from the Hill region (82.21% and 78%, respectively) compared to health workers from the Terai region (80.69% and 51.13%, respectively). Lower knowledge and skill score of service providers from the Terai region where NMR is higher (28 per 1000 live births) than the Hilly region (23 per 1000 live births)(7) is both concerning and unexpected. As such, our study demonstrates the urgency in building and sustaining newborn resuscitation knowledge and skill of health workers from Terai region to have higher impact in reducing NMR in Nepal and eventually achieving SDG target of reducing NMR to 12 per 1000 live births by 2030.

This study is subject to various limitations, and therefore findings should be interpretated cautiously. First, our assessment of facility readiness to provide newborn resuscitation was based on point prevalence. Hence, the study is unable to guarantee continuous availability of essential equipment, supplies, medicines and neonatal resuscitation aids before and after the survey. Second, selection of an equal number of providers per facility irrespective of their scope of service provision or caseload, may have overrepresented providers in smaller facilities while underrepresented providers at larger facilities. Third, the providers may have altered their practice of newborn resuscitation due to the presence of observers. Thus, the actual practice of resuscitation may be underreported due to the Hawthorne effect. Nonetheless, this study offers the most recent information on the competency of Nepalese providers in newborn resuscitation and provides recommendations to further enhance knowledge and skills of service providers.

## Conclusion

This study stresses the need of capacity-building initiatives for newborn resuscitation in response to identified knowledge-skill gaps. These gaps included differences between provinces, facility type, delivery load, and presence of resuscitation aids. As the participation in the LDSC/SSN’ skill retention program was the best predictor of skills, the LDSC/SSN model of newborn resuscitation training, scale up and skill retention and could be one potential cost-effective model to improve the quality of immediate newborn care. We recommend that the MoHP and development partners focus on enhancing newborn resuscitation competency of health workers from the regions that represent the highest burden of neonatal mortality.

## Data Availability

All data underlying the findings are provided as part of the submitted article.

## Acknowledgements

The authors are grateful to all the participating institutions and service providers who participated in the study. We are also thankful to Teresa Gomez, Ph. D for helping in curating geospatial map of the health facilities included in the study. We would like to thank the Church of Jesus Christ of Latter-day Saints for sponsoring LDS Charities, who implemented this program.

## Supporting Information

**S1 Table. Percentage of providers who gave correct response to each item on newborn resuscitation knowledge checklist. *S1_Table*.*docx***

**S2 Table. Percentage of providers who completed specific task correctly during observation of newborn resuscitation. *S2_Table*.*docx***

